# Contactless Depression Screening via Facial Video-derived Heart Rate Variability

**DOI:** 10.1101/2025.05.01.25326621

**Authors:** Min Jhon, Ju-Wan Kim, Kiwook Lee, Dawoon Kim, Se-Hyoun Park, Changheon Kim, Bahngtaik Lim, Seon-Young Kim, Sung-Wan Kim, Jae-Min Kim, Il-Seon Shin, Yoonjoo Choi

## Abstract

**Background:** Depression is a prevalent mental health condition that frequently goes undiagnosed. Heart rate variability (HRV) has emerged as a potential objective marker of depression. Facial video-based HRV measurement offers a novel, contactless approach that could facilitate widespread, non-invasive depression screening.

**Methods:** We analyzed data from 1,453 individuals who completed facial video recordings for HRV analysis and the Patient Health Questionnaire-9 (PHQ-9). A stacking ensemble classifier was developed using HRV features and basic demographic information to classify individuals with depressive symptoms. The ensemble incorporated four base learners (logistic regression, gradient boosting, XGBoost, and SVM) with an SVM meta-learner. Model performance was evaluated using 5-fold cross-validation.

**Results:** The stacking model achieved its best discrimination of AUROC 0.64 (AUPRC 0.45 and MCC 0.21). Incorporating demographic features alongside HRV improved performance over HRV alone. Feature importance analysis revealed that smoking status, sex, and medical comorbidities were the strongest contributors to the predictions.

**Limitations:** The predictive performance was modest, and HRV alone showed limited discrimination. Additionally, the findings are based on a single cohort and require validation in more diverse populations.

**Conclusion:** Facial video-derived HRV, combined with simple demographic factors, can moderately distinguish individuals with depressive symptoms in a contactless manner. Although predictive performance was modest, this non-invasive approach shows promise for accessible, large-scale depression screening.

**Highlights:**

- Facial video-derived HRV enables non-invasive, contactless depression screening.
- Stacking ensemble with SVM meta-learner optimized for MCC in depression screening.
- Combining HRV with demographics improved depression classification vs. HRV alone.
- Moderate yet consistent performance achieved with minimal, non-invasive inputs.

## Introduction

Depression, a common mental disorder, is the leading causes of disability associated with a high level of disease burden. (Collaborators, 2018). Given its high prevalence and significant impacts on individuals and societies, innovative techniques have been developed for the detection of depression. One important aspect of depression detection is that poor health literacy, social stigma, and lack of trust in the therapeutic relationship with professionals are currently recognized as factors that prevent young people from receiving timely mental health treatment (Radez et al., 2021). Consequently, identifying mental health problems using objective data has emerged as a potential solution to these issues. Recently, machine-learning (ML) models have been widely employed to detect depression from objective data such as speech patterns, electroencephalograms, eye movements, social media posts, sleep patterns, activity, and heart rate variability (HRV) (Aldarwish and Ahmad, 2017; Byun et al., 2019; Coutts et al., 2020; Liu et al., 2015; Zhang et al., 2023; Zhu et al., 2020). Among these, HRV has received considerable attention, with systematic reviews confirming an association between HRV and depression (Hartmann et al., 2019).

HRV measures the time variation between heartbeats and serves as a quantitative indicator of autonomic nervous system (ANS) activity (Cygankiewicz and Zareba, 2013). Depressive symptoms are known to be linked to central autonomic network, potentially leading to reduced vagal outflow and altered HRV patterns (Chalmers et al., 2014). Indeed, meta-analyses have consistently shown that individuals diagnosed with major depressive disorder (MDD) tend to exhibit lower HRV compared to non-depressed controls, with the magnitude of HRV reduction often correlating the severity of depression (Kemp et al., 2010; Koch et al., 2019; Wu et al., 2023).

Despite these established associations, the practical utility of HRV for accurately predicting depressive symptoms in diverse populations and real-world settings remains unclear. While recent advances in artificial intelligence (AI) techniques have facilitated more sophisticated analyses, several studies highlight the limitations of relying solely on HRV in depression detection (Byun et al., 2019; Geng et al., 2023; Kim et al., 2017; Sun et al., 2016; Zhang et al., 2011). Early research often suffered from small sample sizes and data collection under controlled conditions, likely limiting the generalizability of findings.

Recent technological progress, especially the advancements of wearable devices and remote photoplethysmography (rPPG), offer a potential solution for collecting HRV data at a larger scale while minimizing participant discomfort and reducing the need for professional administration. However, translating HRV analysis from controlled settings to naturalistic environment present significant challenges. Signals from wearable devices are often susceptible to artifacts from movement and other real-world factors, potentially compromising data quality. Furthermore, studies using wearable-derived HRV data collected have shown mixed results. For example, a recent study analyzing wearable-derived HRV (Hornstein et al., 2022) achieved only limited predictive success, with an area under the receiver operating characteristic curve (AUROC) of 0.56, when classifying participants based on moderate depressive symptoms. Such findings highlight the difficulty of using data from wearable devices for cross-sectional depression prediction amidst numerous real-world sources of variation.

Given these challenges, there is continues interest in exploring novel, accessible and more robust methods for capturing physiological signals related to mental state. Among other detection technologies, measuring HRV using facial images has emerged as a promising approach in this regard (Odinaev et al., 2023; Unursaikhan et al., 2021). This study aimed to investigate the potential of this contactless HRV detection technique based on facial video analysis. We collected facial video data from over 2,000 individuals to derive HRV features, thereby addressing the sample size limitations of previous studies. We then developed and evaluated an ML model using a stacking-based ensemble technique and recursive clustering to handle potentially noisy data, to assess the utility of facial-video-derived HRV in predicting self-reported depressive symptoms. The primary objective was to determine the predictive performance achievable with this approach and explore its potential contribution to the field of objective mental health assessment.

## Materials and Methods

### Study Outline and Participants

This study collected data from August 2, 2021, to October 1, 2023, to investigate mental disorders using a transdiagnostic approach based on common symptoms and processes. The dataset includes scores on psychological scales, interview recordings, HRV, and vital signs such as blood and actigraphy data. All participants provided written consent prior to participation. Key inclusion criteria for the parent study included being > 18 of age being able to wear an actigraphy device and provide digital data such as voice, facial video, smartphone usage. HRV data were collected on-site at the hospital on the day of the participants’ visit. **Figure 1** shows the detailed participant recruitment process and classification flow.

**Figure 1.**
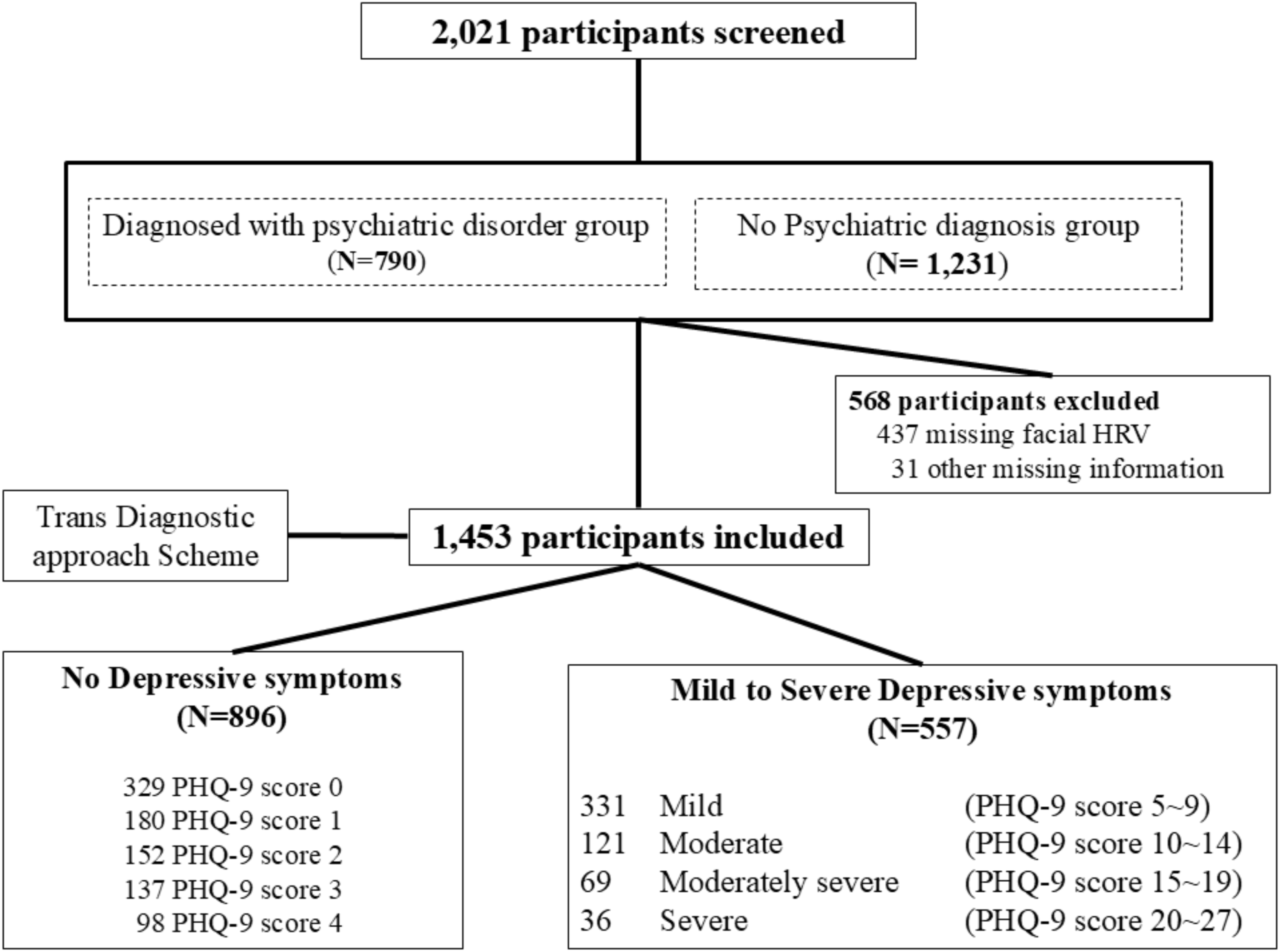
Participant recruitment and classification flow diagram. A total of 2,021 participants were enrolled. After excluding 568 participants due to missing information, 1,453 participants were included for analysis. Participants were classified based on depressive symptom severity measured by the Patient Health Questionnaire-9 (PHQ-9), using a cutoff score of > 4 to define the presence of depressive symptoms. Finally, 559 participants were categorized into the depressive symptoms group, and 894 were categorized as having no depressive symptoms.

The severity of depression was determined by the scale Patient Hospital Questionnaire-9 (PHQ-9) (Kroenke et al., 2001), a widely used and validated tool for screening depression in primary care settings, which assesses the presence and severity of depressive symptoms over the prior 2 weeks. Each item is scored on a scale of 0 to 3, with higher scores indicating more severe symptoms. The total score ranges from 0 to 27. Standard severity categories are: 0–4 (no depressive symptoms), 5–9 (mild), 10–14 (moderate), 15–19 (moderately severe), and 20–27 (severe). In this study, participants were classified using a binary approach based on their PHQ-9. A score of 5 or higher was used to define the presence of depressive symptoms. As an additional measure to evaluate the depressive symptom, the Hospital Anxiety Depression Scale depression subscale (HADS-D) (William, 1976), a self-report measure focusing less on somatic symptoms, was also administered. HADS-D scores were used for descriptive characterization of the sample (**Table 1**) but were not included as outcome measures or predictors in the primary logistic regression or machine learning analyses presented here.

**Table 1.** Characteristics of groups with and without depressive symptoms.

|  | **All**<br>**participants**<br>**(N=1,453)** | **Non-**<br>**depressive**<br>**group**<br>**(N=894)** | **Depressive**<br>**group**<br>**(N=559)** | **Statistical**<br>**coefficient** |
| --- | --- | --- | --- | --- |
| **Sociodemographic characteristics** |  |  |  |  |
| Age, years | 55 (37-66) | 56 (40-66) | 53 (33-64) | U = 228526<br>P = 0.006* |
| Gender, female | 928 (63.9%) | 568 (63.5%) | 360 (64.4%) | $\chi^2 = 0.112$<br>P = 0.738 |
| BMI | 23.9 (21.8-26.3) | 23.8 (21.7-26) | 23.9 (21.8-26.8) | U = 251348<br>P = 0.217 |
| Smoking status, yes | 183 (12.6%) | 94 (10.5%) | 89 (15.9%) | $\chi^2 = 9.133$<br>P = 0.003* |
| Medical comorbidities | 408 (28.1%) | 238 (26.6%) | 170 (30.4%) | $\chi^2 = 2.446$<br>P = 0.118 |
| **Psychiatric assessments** |  |  |  |  |
| PHQ-9 | 3 (1-7) | 1 (0-3) | 9 (6-13) | U = 498967.5<br>P < 0.001* |
| HADS-D | 5 (3-8) | 4 (2-6) | 8 (6-11) | U = 430273.5<br>P < 0.001* |

Values are resented as median (interquartile range) or number (%). Mann-Whitney U-test or the $\chi^2$ test was used.
BMI, Body mass index; PHQ-9, Patient health questionnaire-9; HADS-D, Hospital anxiety depression scale-depression; SD, Standard deviation.
\* P < 0.05

As we adopted a transdiagnostic approach, we categorized groups based solely on the severity of depressive symptoms, regardless of specific diagnoses or treatments. Of the total of 2,021 participants initially enrolled, 1,453 participants were included in the analysis, while 568 were excluded due to missing HRV data. Based on PHQ-9 scores, 559 had mild to severe depressive symptoms and 894 were defined as not being in a depressive state. Written informed consent was collected, and the study was approved by the CNUH and CHUHH Institutional Review Boards (approval nos. CNUH-2021-243, CNUH-2022-216, CNUHH-2021-117, and CNUHH-2022-126).

### Facial video-based HRV detector

Data collection and HRV analysis were completed on the day of each participant’s hospital visit using facial video-based HRV detection software (Korean patent number 10-2150635). Prior to the assessment, participants rested for 5 minutes. They were asked to remove metal accessories, keep their eyes open, and adopt a comfortable sitting position. To minimize biases from movement artifacts, participants were instructed not to move or speak, and to breathe naturally during the test. A standard web camera placed on the monitor captured facial images at 30 frames per second. A well-trained experimenter oversaw all data collection procedures.

The software calculates HRV by remotely sensing heart rate through subtle changes in facial skin color. The region of interest (ROI) on the face is automatically identified. Red, green, and blue (RGB) signals extracted from the ROI are processed to compute a raw PPG signal. This signal is filtered using a Butterworth bandpass filter (0.75–2.5 Hz) to isolate heart rate-related frequency components. Subsequently, the CHROM algorithm, which utilizes specific linear combinations of RGB signals to enhance the pulsatile component while mitigating noise, is applied to derive inter-beat intervals from the filtered PPG signal (Haan and Jeanne, 2013) (**Supplementary Figure S1)**.

Standard time-domain and frequency-domain HRV metrics were then calculated from the interpolated RR intervals following established guidelines (Pham et al., 2021). The validity of this facial video-based HRV estimation approach was examined using the MAHNOB-HCI dataset (Soleymani et al., 2012), which includes synchronized recordings of facial video and ECG from 27 participants under controlled conditions. A comparison between video-derived and ECG-based HRV metrics showed a reasonable level of agreement (**Supplementary Table S1**), supporting the feasibility of this method for non-contact HRV monitoring.

The parameters yielded by the facial video-based HRV detection software and considered in this study are as follows: mean heart rate (HR); standard deviation of the normal-to-normal interval (SDNN); root mean square of successive RR interval differences (RMSSD); percentage of successive RR intervals greater than 50 ms (pNN50); total power (TP); very low frequency (VLF), low frequency (LF), and high frequency (HF) HRV; LF/HF ratio; natural logarithm of TP (LnTP); natural logarithm of VLF (LnVLF); natural logarithm of LF (LnLF); natural logarithm of HF (LnHF); LF divided by the total power [LF(%)]; HF divided by the total power [HF(%)]; and coherence ratio.

### Statistical analyses

Participants were divided into the following two groups according to their PHQ-9 status. Those with no depressive symptoms (PHQ-9 < 5) and those with depressive symptoms (PHQ-9 score of 5 or higher). Sociodemographic characteristics and HRV parameters were compared between the two groups using the Mann-Whitney U-test and the χ^2^ test, as appropriate. For the frequency domain HRV parameters (TP, VLF, LF, HF), natural logarithmic transformations were applied prior to analysis to reduce the skewness.

To examine the association between HRV measures and the presence of depressive symptoms, factors associated with depressive symptoms in univariate analysis (P < 0.05) were entered into logistic regression as independent variables. Demographic factors known to potentially influence HRV and/or depression including age, sex, smoking status (yes/no), the presence of medical comorbidities (yes/no) (defined as a history of one or more of the following conditions: hypertension, diabetes mellitus, angina, or cerebrovascular disease), and body mass index (BMI) were included as covariates.

### Machine learning methodology

A stacking ensemble machine learning model was developed to predict depressive symptom severity using HRV and demographic data. The final stacking ensemble consisted of logistic regression (LR), gradient boosting (GB), extreme gradient boosting (XGB), and support vector machine (SVM) models as base learners. An SVM model was employed as the meta-learner to integrate predictions from the base learners into a final classification decision.

Hyperparameter tuning of each base learner and the final stacking model was performed using the Optuna optimization framework. The optimization objective was set as maximizing the Matthews correlation coefficient (MCC), a robust measure particularly suited for evaluating binary classification performance in imbalanced datasets. Hyperparameter spaces were individually defined for each base model, encompassing parameters such as the regularization strength for LR, learning rates and tree depths for GB and XGB, and kernel parameters for SVM. Optuna (Akiba et al., 2019) was configured to execute a total of 30 optimization trials per model, utilizing the Tree-structured Parzen Estimator (TPE) algorithm (Watanabe, 2023) for efficient hyperparameter exploration.

Model performance and generalization were rigorously assessed using a stratified 5-fold cross-validation procedure. The dataset was divided into five equal subsets, with each subset serving once as a validation set while the remaining four subsets formed the training data. To avoid data leakage and ensure unbiased evaluation, scaling was performed within each training fold independently and subsequently applied to the validation fold. Performance was evaluated by computing a comprehensive set of metrics, including the Matthews correlation coefficient (MCC), area under the receiver operating characteristic curve (AUROC), and area under the precision-recall curve (AUPRC).

Following the training and evaluation of the stacking ensemble, SHapley Additive exPlanations (SHAP) analysis (Lundberg and Lee, 2017) was performed to interpret the model’s predictions. SHAP analysis provided insights into the relative importance and contribution of each feature to the prediction outcomes. SHAP values were computed using the KernelExplainer, leveraging a representative subset of 50 samples from the dataset as background data to enhance computational efficiency. The mean absolute SHAP values across samples were then used to rank the features by importance, allowing clear identification of the most influential demographic and HRV-related predictors of depressive symptoms.

## Results

### Participant Characteristics and Group Differences

A total of 1,453 participants were included in the final analysis, comprising a dataset with a distribution of depressive symptoms broadly consistent with expected prevalence rates (**Table 1** and **Supplementary Figure 2**), with 559 participants (38.5%) classified into the depressive symptom group (PHQ-9 ≥ 5) and 894 participants (61.5%) into the non-depressive group (PHQ-9 < 5). This balanced distribution facilitated robust and meaningful comparative analyses between groups, providing reliable representation for subsequent machine-learning approaches.

The overall median age of participants was 55 years (interquartile range [IQR]: 37–66), and the majority of the sample was female (63.9%). There was a slight yet statistically significant age difference between the groups, with the depressive symptom group being slightly younger (median age: 53 years; IQR: 33–64) than the non-depressive group (median age: 56 years; IQR: 40–66). Nevertheless, this minor difference did not detract from the generally balanced demographic profile between groups.

Sex distribution was generally similar across groups, with females accounting for 64.4% of participants in the depressive symptom group and 63.5% in the non-depressive group. Body mass index (BMI) also showed high comparability, with median values essentially identical (23.9 for both groups). Smoking status emerged as significantly different, being more common among participants with depressive symptoms (15.9%) than those without (10.5%; p = 0.003, χ² test). In contrast, medical comorbidity rates, defined as a history of hypertension, diabetes mellitus, angina or cerebrovascular disease, were well-matched and balanced between groups (30.4% and 26.6% respectively; p = 0.118, χ² test). Hence, aside from smoking, the primary health variables showed balanced distributions across both groups.

An independent measure of depression severity with the HADS-D shows a good correlation with PHQ-9. Participants in the depressive symptom group exhibited significantly higher HADS-D scores (median: 8, IQR: 6–11) compared to those in the non-depressive group (median: 4, IQR: 2–6; p < 0.001, Mann-Whitney U test). This strong and significant difference in HADS-D scores shows the appropriateness and robustness of the initial classification criteria, indicating that the PHQ-9-based grouping accurately reflects meaningful variations in depressive symptomatology.

### Associations Between HRV Parameters and Depressive Symptoms

Univariate comparisons of HRV measures between these groups revealed several significant differences (**Table 2** and **Supplementary Figure S3**). Participants with depressive symptoms exhibited a higher mean resting heart rate compared to non-depressive counterparts (median: 75.4 bpm vs. 73.9 bpm; p < 0.001). In the time-domain HRV analyses, individuals in the depressive group displayed reduced HRV, with lower SDNN and RMSSD values (both p < 0.01). However, the pNN50 did not differ significantly between groups (p = 0.071). Analysis of frequency-domain HRV indices also indicated that participants with depressive symptoms had significantly reduced HRV across multiple measures. The depressive group exhibited lower total power (LnTP), as well as reduced values across specific frequency bands including LnVLF, LnLF, and LnHF (all p ≤ 0.001), whereas the LF/HF ratio did not differ significantly (p = 0.112). Collectively, these results suggest that depressive symptoms are consistently associated with an increased resting heart rate and broadly suppressed HRV.

**Table 2.** Heart rate variability measures of groups with and without depressive symptoms.

|  | **All**<br>**participants**<br>**(N=1,453)** | **Non-**<br>**depressive**<br>**group**<br>**(N=894)** | **Depressive**<br>**group**<br>**(N=559)** | **Statistical**<br>**coefficient** |
| --- | --- | --- | --- | --- |
| Mean HR, bpm | 74.5 (68.7-80.4) | 73.9 (68-79.5) | 75.4 (70.4-81.7) | U = 285146<br>P < 0.001* |
| SDNN, ms | 45.6 (38.3-15.4) | 46.2 (39.8-54.2) | 44.2 (36.6-52.1) | U = 222299<br>P < 0.001* |
| RMSSD, ms | 56.2 (47.6-66.9) | 57.1 (48.5-67.4) | 54.6 (46-65.6) | U = 226109<br>P = 0.002* |
| PNN50, % | 23.9 (19.8-27.7) | 24.1 (20.4-27.7) | 23.6 (18.9-27.8) | U = 235832<br>P = 0.071 |
| LnTP, ms<sup>2</sup> | 7.29 (6.94-7.67) | 7.34 (7.02-7.71) | 7.24 (6.8-7.63) | U = 220184<br>P < 0.001* |
| LnVLF, ms<sup>2</sup> | 5.22 (4.68-5.22) | 5.29 (4.71-5.75) | 5.12 (4.63-5.64) | U = 224758<br>P = 0.001* |
| LnLF, ms<sup>2</sup> | 6.13 (5.67-6.53) | 6.17 (5.75-6.58) | 6.06 (5.57-6.45) | U = 219530<br>P < 0.001* |
| LnHF, ms<sup>2</sup> | 6.68 (6.26-7.06) | 6.72 (6.34-7.1) | 6.6 (6.18-7.02) | U = 224875<br>P = 0.001* |
| LF/HF ratio | 0.57 (0.45-0.73) | 0.58 (0.45-0.74) | 0.56 (0.45-0.70) | U = 237508<br>P = 0.112 |
| Coherence ratio | 0.02 (0.02-0.04) | 0.02 (0.01-0.04) | 0.02 (0.02-0.04) | U = 254879<br>P = 0.512 |

Values are presented as median (interquartile range) or number (%). Mann-Whitney U-test or the $\chi^2$ test was used.
HR, Heart rate; SDNN, Standard deviation of the normal-to-normal intervals; RMSSD, Root mean square of successive RR interval differences; pNN50, Percentage of successive RR intervals greater than 50 ms; LF/HF, Low-frequency to high-frequency ratio; LnTP, Natural logarithm of total power; LnVLF, Natural logarithm of very low-frequency; LnLF, Natural logarithm of low-frequency; LnHF, Natural logarithm of high-frequency; SD, Standard deviation.
\* P < 0.05

**Table 3.** Multivariate analyses of heart rate variability measures associated with depressive symptoms.

|  | **Adjusted OR<sup>a</sup>** | **95% CI** | **p-value** |
| --- | --- | --- | --- |
| Mean HR, bpm | 1.023 | 1.010 – 1.036 | <0.001* |
| SDNN, ms | 0.987 | 0.977 – 0.997 | 0.009* |
| RMSSD, ms | 0.992 | 0.984 – 1.000 | 0.038 |
| LnTP, ms<sup>2</sup> | 0.757 | 0.627 – 0.915 | 0.004* |
| LnVLF, ms<sup>2</sup> | 0.831 | 0.726 – 0.953 | 0.008* |
| LnLF, ms<sup>2</sup> | 0.748 | 0.630 – 0.888 | 0.001* |
| LnHF, ms<sup>2</sup> | 0.809 | 0.676 – 967 | 0.020* |

HR, Heart rate; SDNN, Standard deviation of the normal-to-normal intervals; RMSSD, Root mean square of successive RR interval differences; LnTP, Natural logarithm of total power; LnVLF, Natural logarithm of very low-frequency; LnLF, Natural logarithm of low-frequency; LnHF, Natural logarithm of high-frequency; OR, Odds ratio; CI, Confidence interval.
<sup>a</sup> Adjusted for age, sex, smoking status, medical illness status and body mass index.
\* P < 0.05

To determine which HRV measures were independently associated with depressive symptoms, we conducted multivariable logistic regression analyses adjusting for key demographic and clinical covariates, including age, sex, BMI, smoking status, and medical comorbidities (**Supplementary Table S2**). Results from these adjusted analyses confirmed that mean heart rate remained a significant independent predictor of depressive symptoms, with each 1 bpm increase in mean heart rate associated with a 2.3% greater odds of experiencing depressive symptoms (adjusted odds ratio (OR): 1.023; 95% confidence interval (CI): 1.010– 1.036; p < 0.001). Higher HRV values also demonstrated protective associations with depressive symptoms. Increased SDNN and RMSSD were independently linked to lower odds of depressive symptoms, as were higher values in frequency-domain parameters such as LnTP, LnVLF, LnLF, and LnHF (all p < 0.05).

These robust multivariable findings reinforce the role of reduced autonomic flexibility, as indicated by lowered HRV and elevated resting heart rate, in the presentation of depressive symptoms. The combined univariate and adjusted analyses indicate the strength and consistency of associations between specific HRV parameters and depressive symptomatology.

### Stacking Ensemble Model for the Prediction of Depression

We developed a machine learning model to classify depressive symptom status using a stacking ensemble framework. The final ensemble integrated four diverse base learners: logistic regression (LR), gradient boosting (GB), extreme gradient boosting (XGB), and support vector machine (SVM), with an SVM model serving as the meta-learner. This design allowed the meta-learner to combine the strengths of each algorithm’s predictions into a superior final decision. Although a random forest (RF) classifier was initially considered, it was excluded from the final stack due to its comparatively low individual performance (**Figure 2A**). To robustly evaluate model performance, we employed a stratified 5-fold cross-validation strategy.

**Figure 2.**
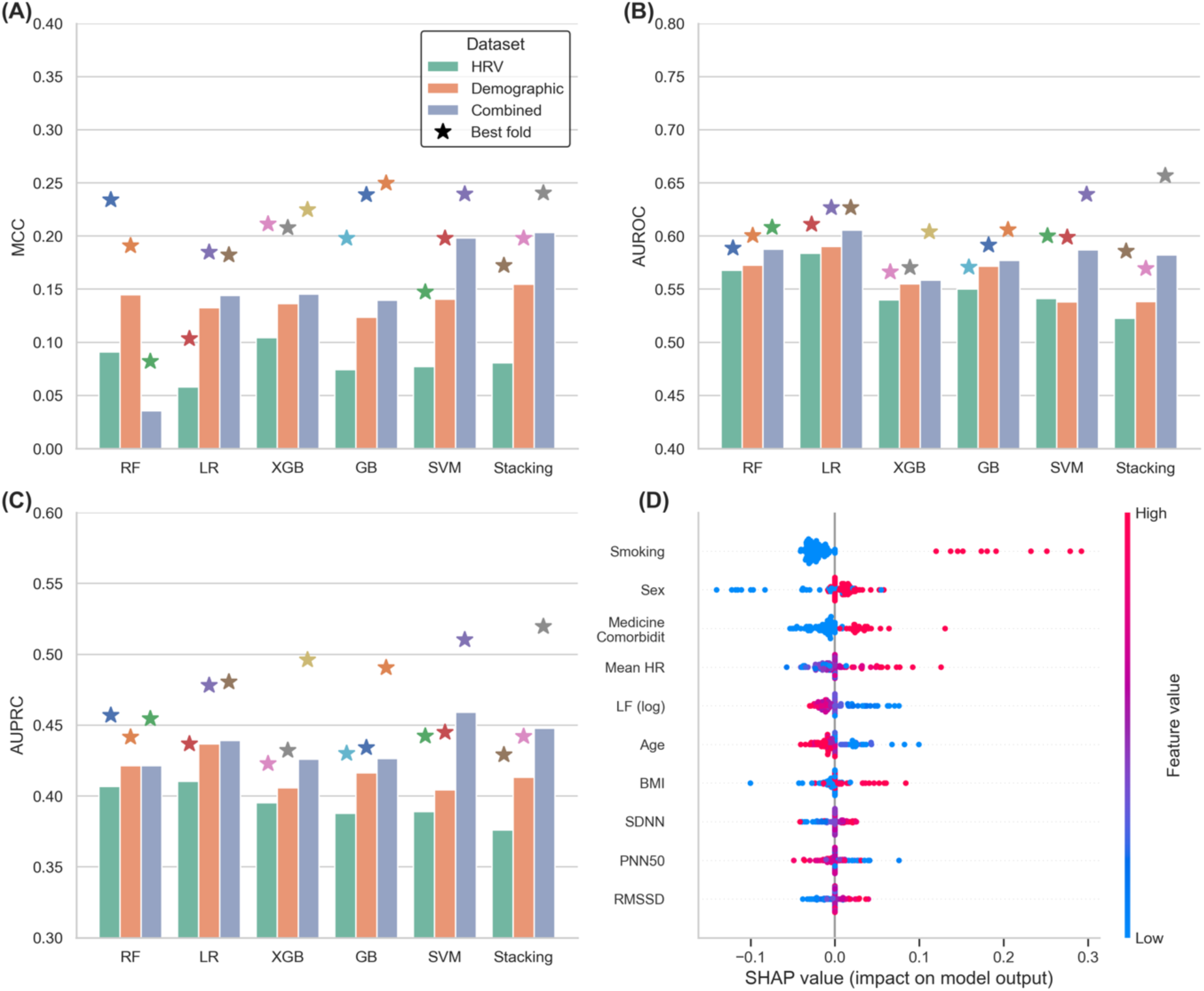
Performance metrics of the stacking ensemble model predicting depressive symptoms. (A) Comparison of Matthews correlation coefficient (MCC), (B) area under the receiver operating characteristic curve (AUROC), and (C) area under the precision-recall curve (AUPRC) for three distinct feature sets: heart rate variability (HRV) alone, demographic information alone, and combined (HRV + demographic). The stacking ensemble model used logistic regression (LR), gradient boosting (GB), extreme gradient boosting (XGB), and support vector machine (SVM) as base learners, with SVM as the meta-learner. Hyperparameters were optimized via Optuna using MCC as the optimization criterion. The stacking ensemble yielded consistently higher MCC values across all test sets. While AUROC and AUPRC values for the stacking ensemble were not always highest in every fold, the ensemble’s best-performing fold consistently outperformed individual models. (D) The SHAP analysis shows that the demographic information is the most impactful features while some HRV features also contribute to the depression prediction.

Optimal hyperparameters for each base model and the meta-learner were determined using Optuna with careful attention to the class imbalance in our dataset. Importantly, we set the optimization objective to maximize the MCC rather than AUROC. This choice was made after observing that tuning for AUROC would simply predict every case as non-depressed (majority class). As a result, the final tuned ensemble was better calibrated to detect depressive symptoms, avoiding the trivial all-negative prediction strategy. All model training and validation processes were confined within each cross-validation fold to prevent any information leakage (e.g., data scaling was fit on each training fold and applied to its respective test fold).

We explored the model’s performance using three different feature sets (HRV-only, demographic-only, and a combined feature set) (**Figure 2**). Strikingly, models trained solely on HRV features had very limited predictive power. HRV alone was not very informative for distinguishing those with depressive symptoms from those without. In contrast, using demographic features only yielded stronger predictive performance than HRV alone, indicating that these personal factors are highly relevant to depression status. Of the three, the combined feature set (HRV + demographics) performed the best, suggesting that while HRV metrics by themselves provide limited signals, they do add some complementary information when combined with demographic data. In other words, demographic factors carried the bulk of predictive information, but incorporating physiological HRV measures yielded a slight yet meaningful boost in model effectiveness.

The stacking ensemble approach proved to be the top-performing model in our evaluations, albeit by a small margin. As shown in **Figure 2A**, the ensemble’s MCC was consistently the highest across all five test folds, outperforming each individual base learner’s MCC on every fold by a slight amount. This consistency indicates that the ensemble approach improved the reliability of predictions for the minority class (depressive cases) in each split. Notably, the improvement in MCC was only marginal, but it was systematically in favor of the stacking model for every test subset.

In terms of overall discrimination, the stacking model achieved competitive results: its AUROC and AUPRC were not the highest on average (**Figure 2B and 2C**), yet the best fold performance of the ensemble in both AUROC and AUPRC was still higher than that of any single model. These findings demonstrate that the ensemble approach, even if only incrementally better on certain metrics, provided a robust and generalizable solution for predicting depressive symptoms from the feature set.

### Feature Importance Analysis

To understand the model’s predictions, we performed a post-hoc interpretability analysis using SHAP (SHapley Additive exPlanations) values. Examining the final stacking ensemble’s feature importances revealed a mix of demographic and HRV features among the top predictors. In fact, about half of the top 10 features ranked by absolute SHAP impact were HRV-derived metrics and the other half were demographic variables (**Figure 2D**).

Crucially, however, the top three features with the highest influence on the model were all demographic factors: smoking status, sex, and medical comorbidities. We also inspected the feature importance profiles of the individual base learners for comparison (**Supplementary Figure S4**). Interestingly, most models relied extremely heavily on the smoking feature when making their predictions. In the GB, XGB, and SVM classifiers, smoking status dominated the feature importance rankings, often far exceeding the contribution of any other feature. The stacking meta-learner, which effectively averages across these, likewise reflected this dominance in the overall top features as noted above. The exception was the LR model. In LR’s case, features like sex, age, or certain HRV parameters received more balanced weights alongside smoking. This difference likely arises because logistic regression, being a simpler linear model, had to assign coefficients to all inputs simultaneously, whereas tree-based models (GB/XGB) and SVM could concentrate their decision boundaries on one highly predictive attribute (smoking) before considering others. These findings contribute to the growing evidence that combining physiological signals with personal context is a promising direction for improving automated depression screening tools.

## Discussion

This study provides one of the first large-scale evaluations of contactless, facial video-based HRV as a potential tool for depression screening in naturalistic clinical settings. Using a sample of 1,453 participants, we demonstrated that while traditional demographic factors such as smoking status, sex, and medical comorbidities remain dominant predictors of depressive symptoms. Facial-video-derived HRV features contributed additional predictive value. Specifically, smoking status, sex, and medical comorbidities had greater predictive influence than any individual HRV metric. Previous studies also showed that smokers have roughly twice the risk of developing depression compared to nonsmokers (Hu et al., 2025), and female sex is a recognized risk factor, with depression about 50% more common in women than in men (Albert, 2015). Likewise, the presence of medical comorbidities, often correlates with chronic stress and increased depression risk (Benton et al., 2007). From a sampling perspective, it is possible that these demographic variables explained much of the variance in our cohort’s depression status, making them easy targets for the machine learning model to leverage. In essence, our findings reaffirm that health habits can be as telling as physiological signals when assessing depression risk.

While HRV features alone were relatively weak predictors in our dataset, they did show expected patterns and added value when combined with demographics. Consistent with prior literature, participants with depressive symptoms tended to have reduced HRV. For instance, lower time-domain measures (SDNN, RMSSD) and diminished power in various frequency bands reflect autonomic nervous system dysregulation (Hartmann et al., 2019). Models using only HRV features yielded modest discrimination as reported in a recent study with AUROC often in the 0.6 range (Eriksson et al., 2024). As HRV is influenced by many factors besides mood, though real, is subtle and variable across individuals.

The encouraging finding is that when we integrated HRV with the dominant demographic features, the model’s performance improved consistently. The inclusion of HRV features provided a modest boost over demographics alone, suggesting that HRV does capture a piece of the depression phenotype that demographics do not. In practical terms, HRV served as a complementary biomarker: by adding information about autonomic state, it helped the model pick up on depression cases that might not be fully explained by age, sex, or lifestyle factors. Depression is associated with both genetic vulnerability and environmental factors. Therefore, the use of a single assessment tool for diagnosis may be inappropriate (Kwong et al., 2019; Saveanu and Nemeroff, 2012). Although many studies have described correlations between depression and various biomarkers, such associations have not been fully incorporated into clinical practice.

Our stacking ensemble, which was optimized for the MCC, demonstrated only modest overall accuracy but proved to be robust and consistent in its predictions. The best ensemble model achieved a balanced performance (for example, an MCC around 0.21 with an AUROC of about 0.64 in our sample), indicating moderate ability to distinguish depressive from non-depressive cases. Although these metrics are not high in an absolute sense, the ensemble’s strength lay in how reliably it performed across different threshold definitions and subsamples.

It is worth emphasizing that the simplicity and non-invasiveness of our input features make the obtained performance more acceptable from a practical standpoint. Our method, requiring only a brief facial video and basic demographic information via a smartphone or laptop camera, offers a highly deployable solution for real-world settings. Given that many conventional depression screening tools require people to self-report their mood or involve resource-intensive clinical interviews, a camera-based physiological screening combined with demographics has advantages in situations where those traditional approaches are not feasible or are avoided due to stigma.

Despite these strengths, there are several limitations in this study. First, although facial video-derived HRV provides a non-contact and scalable measurement, it remains more susceptible to environmental artifacts, such as lighting variations and participant movement. Second, depressive symptom classification primarily relied on self-reported PHQ-9 scores, which may not fully correspond to clinical diagnoses. However, we additionally administered the HADS-D to enhance the reliability of depressive symptom assessment. Third, our sample primarily consisted of participants recruited from clinical settings in South Korea. Future studies should aim to validate the model across more diverse and representative populations to ensure broader generalizability.

While our focus was on screening for current depressive symptoms, future work could integrate this tool into a stepped-care model – using contactless HRV screening as an initial filter, followed by more definitive assessment for those flagged. Although our current model has only modest accuracy, it establishes a foundation for accessible depression screening. With further refinement such as adding complementary data streams, expanding to larger and more varied samples, and leveraging more sophisticated features, we anticipate that the performance can be improved. Our findings reinforce that even a quick, contactless assessment can glean meaningful insights into mental health, and they encourage continued innovation at the intersection of digital biomarkers and psychiatry.

## Conclusion

This study demonstrates the feasibility of using contactless, facial video-based HRV measurement combined with basic demographic information for preliminary depression screening. Although demographic factors such as smoking status, sex, and medical comorbidities were the strongest predictors, the inclusion of HRV features modestly improved classification performance. A stacking ensemble model optimized for the MCC achieved consistent and balanced discrimination across validation folds. Given the simplicity and scalability of our method, it holds promise as an accessible tool for large-scale mental health screening, with future work needed to improve performance through additional data integration and model refinement.

## Statement of Ethics

All patients gave written informed consent to participate in the study and use their data. The study was conducted in accordance with the Helsinki Declaration of 1975, as revised in 2008 and approved by the Ethics Commission of the Chonnam National University Hopital Institutional Review Board (CNUH-2021-243, CNUH-2022-216) and the Chonnam National University Hwasun Hospital Institutional Review Board (CNUHH-2021-117, and CNUHH-2022-126) as it uses de-identified data.

## CRediT authorship contribution statement

**Min Jhon**: Study design, Conceptualization, Writing – original draft, review & editing. **Ju-Wan Kim**: Study design, Conceptualization, Writing – original draft, review & editing. **Kiwook Lee**: Conceptualization, Formal analysis, Writing – original draft. **Dawoon Kim**: Conceptualization, Formal analysis. **Se-Hyoun Park**: Data curation, Validation. **Changheon Kim**: Data curation, Formal analysis. **Bahngtaik Lim**: Data curation, Validation. **Seon-Yong Kim**: Data curation, Validation. **Sung-Wan Kim**: Data curation, Validation. **Jae-Min Kim**: Conceptualization, Validation, Writing – review & editing. **Il-Seon Shin**: Conceptualization, Validation, Writing – review & editing. **Yoonjoo Choi**: Study design, Conceptualization, Writing – original draft, review & editing.

## Data Availability

All data produced in the present study are available upon reasonable request to the authors.

## Acknowledgments

The authors are grateful to the staff of the CNUH and CNUHH psychiatry research teams for assisting in the collection of patient data, and to Dr. Kim Jae Chang and Dr. Kim Sun Hee for providing the space for data analysis and data collection.

## References

1. Akiba, T., Sano, S., Yanase, T., Ohta, T., Koyama, M., 2019. Optuna: A Next-generation Hyperparameter Optimization Framework.

2. Albert, P.R., 2015. Why is depression more prevalent in women? Journal of Psychiatry and Neuroscience. 40, 219–221. 10.1503/jpn.150205.

3. Aldarwish, M.M., Ahmad, H.F., 2017. Predicting depression levels using social media posts, 2017 IEEE 13th international Symposium on Autonomous decentralized system (ISADS). IEEE, pp. 277–280.

4. Benton, T., Staab, J., Evans, D.L., 2007. Medical co-morbidity in depressive disorders. Annals of Clinical Psychiatry. 19, 289–303.

5. Byun, S., Kim, A.Y., Jang, E.H., Kim, S., Choi, K.W., Yu, H.Y., Jeon, H.J., 2019. Detection of major depressive disorder from linear and nonlinear heart rate variability features during mental task protocol. Computers in biology and medicine. 112, 103381.

6. Chalmers, J.A., Quintana, D.S., Abbott, M.J.-A., Kemp, A.H., 2014. Anxiety disorders are associated with reduced heart rate variability: a meta-analysis. Frontiers in psychiatry. 5, 80.

7. Collaborators, G.D.a.I.i.a.P., 2018. Global, regional, and national incidence, prevalence, and years lived with disability for 354 diseases and injuries for 195 countries and territories, 1990-2017: a systematic analysis for the Global Burden of Disease Study 2017. Lancet (London, England). 392, 1789–1858. 10.1016/s0140-6736(18)32279-7.

8. Coutts, L.V., Plans, D., Brown, A.W., Collomosse, J., 2020. Deep learning with wearable based heart rate variability for prediction of mental and general health. Journal of Biomedical Informatics. 112, 103610.

9. Cygankiewicz, I., Zareba, W., 2013. Autonomic Nervous System: Chapter 31. Heart rate variability. Elsevier Inc. Chapters.

10. Eriksson, A., Kimmel, M.C., Furmark, T., Wikman, A., Grueschow, M., Skalkidou, A., Frick, A., Fransson, E., 2024. Investigating heart rate variability measures during pregnancy as predictors of postpartum depression and anxiety: an exploratory study. Translational psychiatry. 14, 203. 10.1038/s41398-024-02909-9.

11. Geng, D., An, Q., Fu, Z., Wang, C., An, H., 2023. Identification of major depression patients using machine learning models based on heart rate variability during sleep stages for pre-hospital screening. Comput Biol Med. 162, 107060. 10.1016/j.compbiomed.2023.107060.

12. Haan, G.d., Jeanne, V., 2013. Robust Pulse Rate From Chrominance-Based rPPG. IEEE Transactions on Biomedical Engineering. 60, 2878–2886. 10.1109/TBME.2013.2266196.

13. Hartmann, R., Schmidt, F.M., Sander, C., Hegerl, U., 2019. Heart rate variability as indicator of clinical state in depression. Frontiers in psychiatry. 9, 735.

14. Hornstein, S., Seiler, M., Hoffman, V., Nelson, B., Aschbacher, K., Ritter, K., Hilbert, K., 2022. Association of Depressive Symptoms with Resting Heart Rate Variability recorded from a Wearable Device under Naturalistic Conditions: A Machine Learning Study. PsyArXiv. 10.31234/osf.io/9z3pr.

15. Hu, Z., Cui, E., Chen, B., Zhang, M., 2025. Association between cigarette smoking and the risk of major psychiatric disorders: a systematic review and meta-analysis in depression, schizophrenia, and bipolar disorder. Frontiers in medicine. 12, 1529191. 10.3389/fmed.2025.1529191.

16. Kemp, A.H., Quintana, D.S., Gray, M.A., Felmingham, K.L., Brown, K., Gatt, J.M., 2010. Impact of depression and antidepressant treatment on heart rate variability: a review and meta-analysis. Biological psychiatry. 67, 1067–1074.

17. Kim, E.Y., Lee, M.Y., Kim, S.H., Ha, K., Kim, K.P., Ahn, Y.M., 2017. Diagnosis of major depressive disorder by combining multimodal information from heart rate dynamics and serum proteomics using machine-learning algorithm. Progress in neuro-psychopharmacology & biological psychiatry. 76, 65–71. 10.1016/j.pnpbp.2017.02.014.

18. Koch, C., Wilhelm, M., Salzmann, S., Rief, W., Euteneuer, F., 2019. A meta-analysis of heart rate variability in major depression. Psychological medicine. 49, 1948–1957.

19. Kroenke, K., Spitzer, R.L., Williams, J.B., 2001. The PHQ-9: validity of a brief depression severity measure. Journal of general internal medicine. 16, 606–613. 10.1046/j.1525-1497.2001.016009606.x.

20. Kwong, A.S., López-López, J.A., Hammerton, G., Manley, D., Timpson, N.J., Leckie, G., Pearson, R.M., 2019. Genetic and environmental risk factors associated with trajectories of depression symptoms from adolescence to young adulthood. JAMA Network Open. 2, e196587–e196587.

21. Liu, Z., Hu, B., Yan, L., Wang, T., Liu, F., Li, X., Kang, H., 2015. Detection of depression in speech, 2015 international conference on affective computing and intelligent interaction (ACII). IEEE, pp. 743–747.

22. Lundberg, S.M., Lee, S.-I., 2017. A unified approach to interpreting model predictions, Proceedings of the 31^st^ International Conference on Neural Information Processing Systems. Curran Associates Inc., Long Beach, California, USA, pp. 4768–4777.

23. Odinaev, I., Wong, K.L., Chin, J.W., Goyal, R., Chan, T.T., So, R.H.Y., 2023. Robust Heart Rate Variability Measurement from Facial Videos. Bioengineering (Basel, Switzerland). 10. 10.3390/bioengineering10070851.

24. Pham, T., Lau, Z.J., Chen, S.H.A., Makowski, D., 2021. Heart Rate Variability in Psychology: A Review of HRV Indices and an Analysis Tutorial. Sensors. 21, 3998.

25. Radez, J., Reardon, T., Creswell, C., Orchard, F., Waite, P., 2021. Adolescents’ perceived barriers and facilitators to seeking and accessing professional help for anxiety and depressive disorders: a qualitative interview study. European child & adolescent psychiatry. 1–17.

26. Saveanu, R.V., Nemeroff, C.B., 2012. Etiology of depression: genetic and environmental factors. Psychiatric clinics. 35, 51–71.

27. Soleymani, M., Lichtenauer, J., Pun, T., Pantic, M., 2012. A Multimodal Database for Affect Recognition and Implicit Tagging. IEEE Transactions on Affective Computing. 3, 42–55. 10.1109/T-AFFC.2011.25.

28. Sun, G., Shinba, T., Kirimoto, T., Matsui, T., 2016. An Objective Screening Method for Major Depressive Disorder Using Logistic Regression Analysis of Heart Rate Variability Data Obtained in a Mental Task Paradigm. Front Psychiatry. 7, 180. 10.3389/fpsyt.2016.00180.

29. Unursaikhan, B., Tanaka, N., Sun, G., Watanabe, S., Yoshii, M., Funahashi, K., Sekimoto, F., Hayashibara, F., Yoshizawa, Y., Choimaa, L., Matsui, T., 2021. Development of a Novel Web Camera-Based Contact-Free Major Depressive Disorder Screening System Using Autonomic Nervous Responses Induced by a Mental Task and Its Clinical Application. Frontiers in physiology. 12, 642986. 10.3389/fphys.2021.642986.

30. Watanabe, S., 2023. Tree-Structured Parzen Estimator: Understanding Its Algorithm Components and Their Roles for Better Empirical Performance.

31. William, G., 1976. ECDEU assessment manual for psychopharmacology. US Department of Health, Education, and Welfare, Public Health Service, Alcohol, Drug Abuse, and Mental Health Administration, National Institute of Mental Health, Psychopharmacology Research Branch, Division of Extramural Research Programs.

32. Wu, Q., Miao, X., Cao, Y., Chi, A., Xiao, T., 2023. Heart rate variability status at rest in adult depressed patients: a systematic review and meta-analysis. Frontiers in Public Health. 11, 1243213.

33. Zhang, D., Qu, Y., Zhai, S., Li, T., Xie, Y., Tao, S., Zou, L., Tao, F., Wu, X., 2023. Association between healthy sleep patterns and depressive trajectories among college students: a prospective cohort study. Bmc Psychiatry. 23, 182.

34. Zhang, Z.-X., Tian, X.-W., Lim, J., 2011. New algorithm for the depression diagnosis using HRV: A neuro-fuzzy approach. Proceedings of 2011 International Symposium on Bioelectronics and Bioinformatics, ISBB 2011. 10.1109/ISBB.2011.6107702.

35. Zhu, J., Wang, Z., Gong, T., Zeng, S., Li, X., Hu, B., Li, J., Sun, S., Zhang, L., 2020. An improved classification model for depression detection using EEG and eye tracking data. IEEE transactions on nanobioscience. 19, 527–537.

