## Supplementary Figure S1 for "Contactless Depression Screening via Facial Video-derived Heart Rate Variability"

**Supplementary Figure 1. Facial video-based heart rate variability (HRV) detection.** (A) A camera captures a facial video of the subject, and the software automatically identifies and adjusts the position and size of the region of interest (ROI) around the face. (B) RGB (Red, Green, and Blue) signals are extracted from the facial skin within the ROI for each frame (captured at 30 frames per second). (C) The raw RGB signals are processed through bandpass filtering (Butterworth filter, 0.75–2.5 Hz) to generate a filtered remote photoplethysmography (rPPG) signal. Peak detection is then performed on this filtered signal to calculate inter beat intervals (RR intervals) based on the frame rate, from which heart rate and variability metrics can be determined.

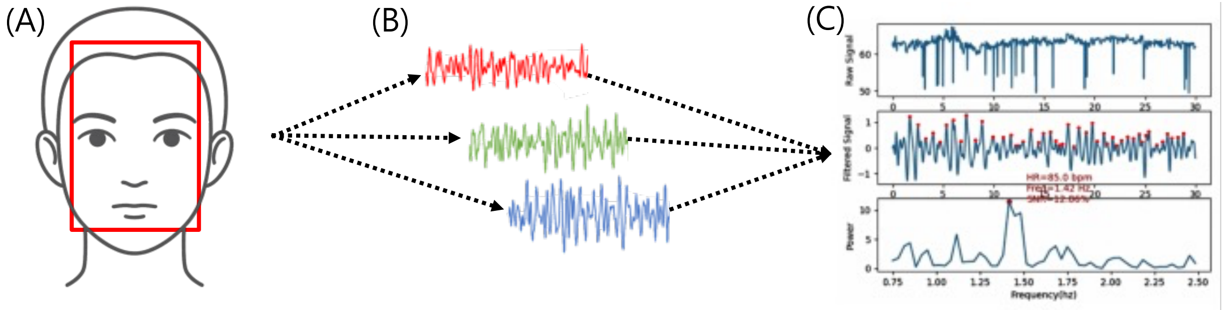

**Supplementary Figure S2. Demographic characteristics and psychological scale scores of participants.** The figure illustrates the distribution of key demographic and psychological variables between depressive and non-depressive groups. Depression status was determined using the Patient Health Questionnaire-9 (PHQ-9) with a cutoff score of ≥5. The horizontal dashed lines indicate median values calculated from the total participant group (black outline). Box plots overlaid on violin plots show the median and interquartile ranges. Individual data points are displayed as small dots. The stacked bar plot shows percentages for categorical demographic variables (sex, smoking status, and medicine use).

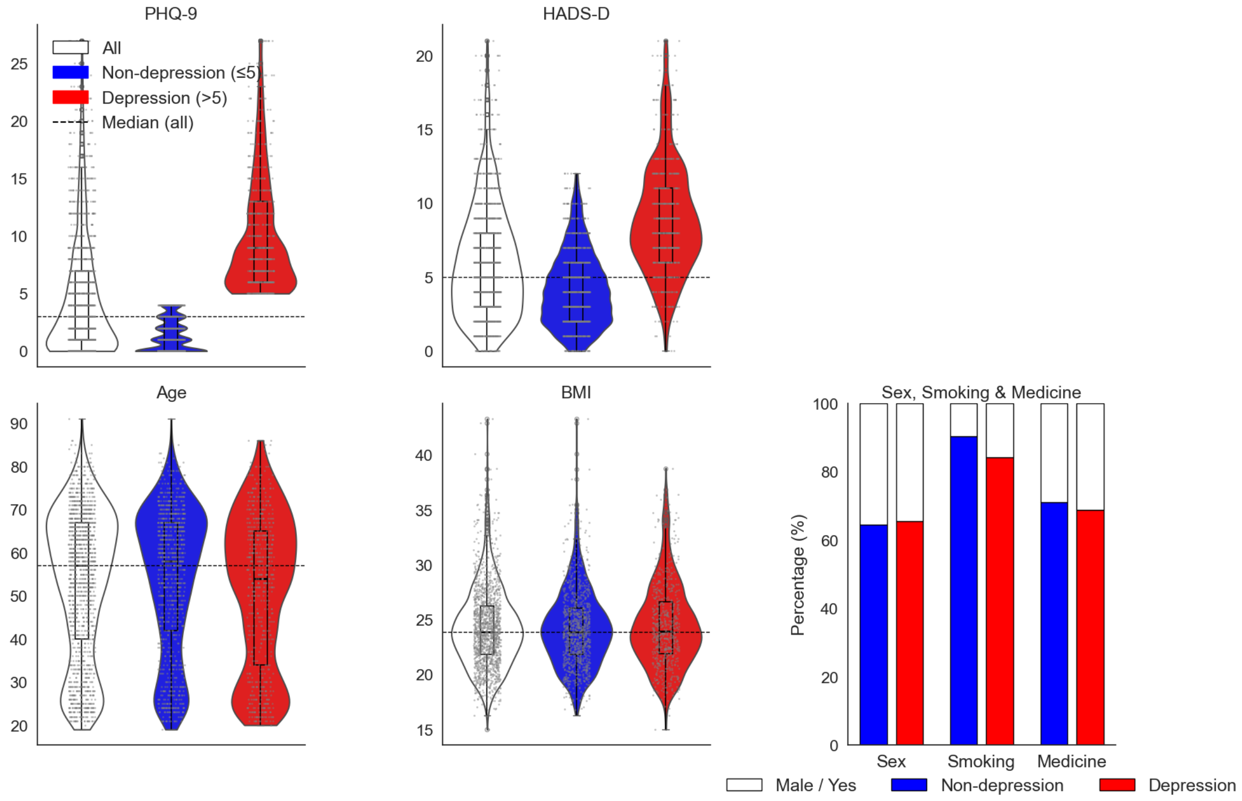

**Supplementary Figure S3. Distribution of heart rate variability (HRV) metrics among participants.** This figure depicts the distribution of individual HRV metrics used in the machine learning analyses, compared between depressive (red; PHQ-9 ≥ 5) and non-depressive (blue; PHQ-9 < 5) groups.

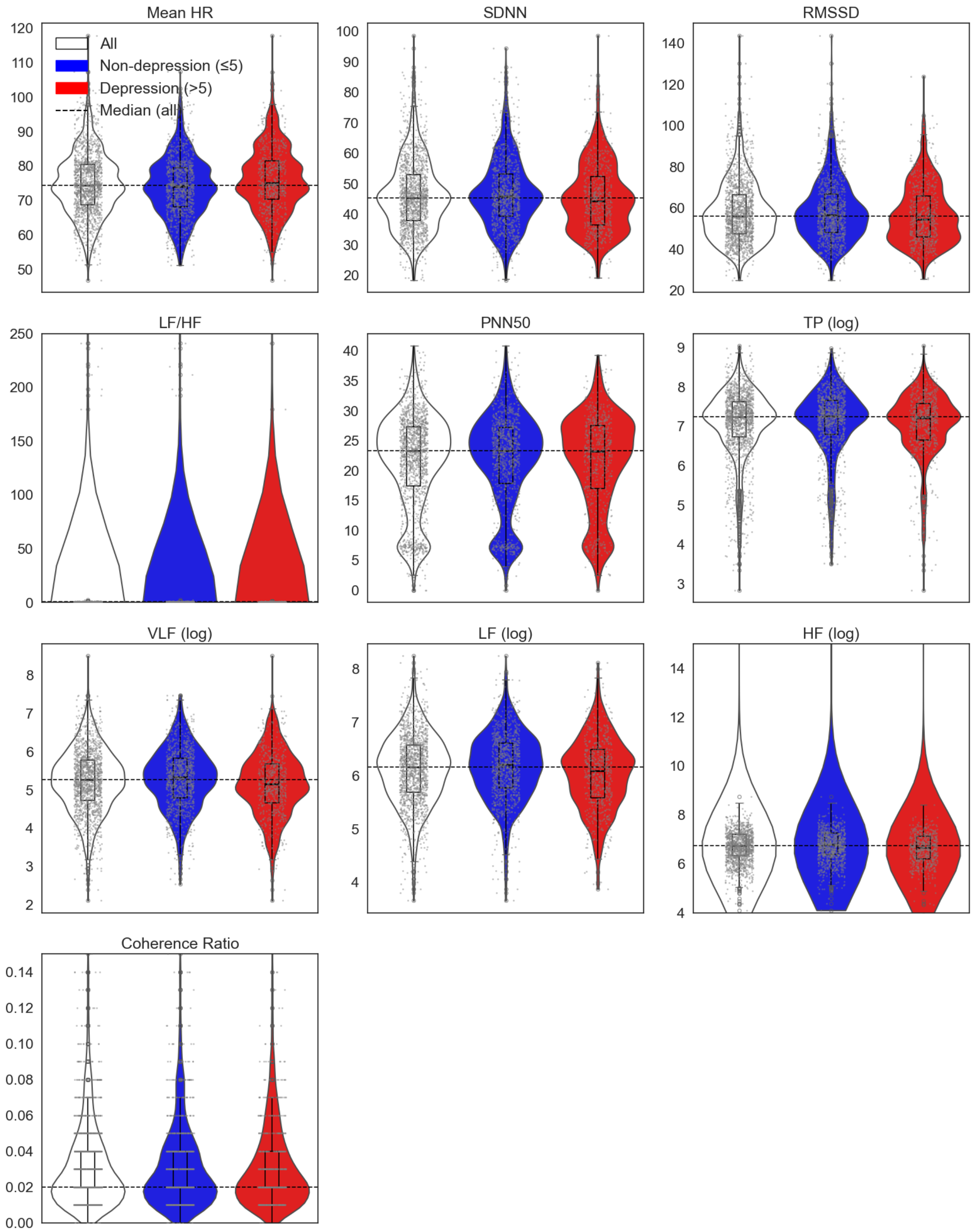

**Supplmentary Figure S4. SHAP feature importance for individual base learner models.** Feature importance analyses were conducted using SHapley Additive exPlanations (SHAP) values to identify and rank features by their contributions to the model predictions for each of the individual base learners, (A) LR, (B) GB, (C) XGB, and (D) SVM. The top ten features identified for each model included a balanced mix of demographic variables and heart rate variability (HRV) metrics, though demographic features dominated the highest rankings.

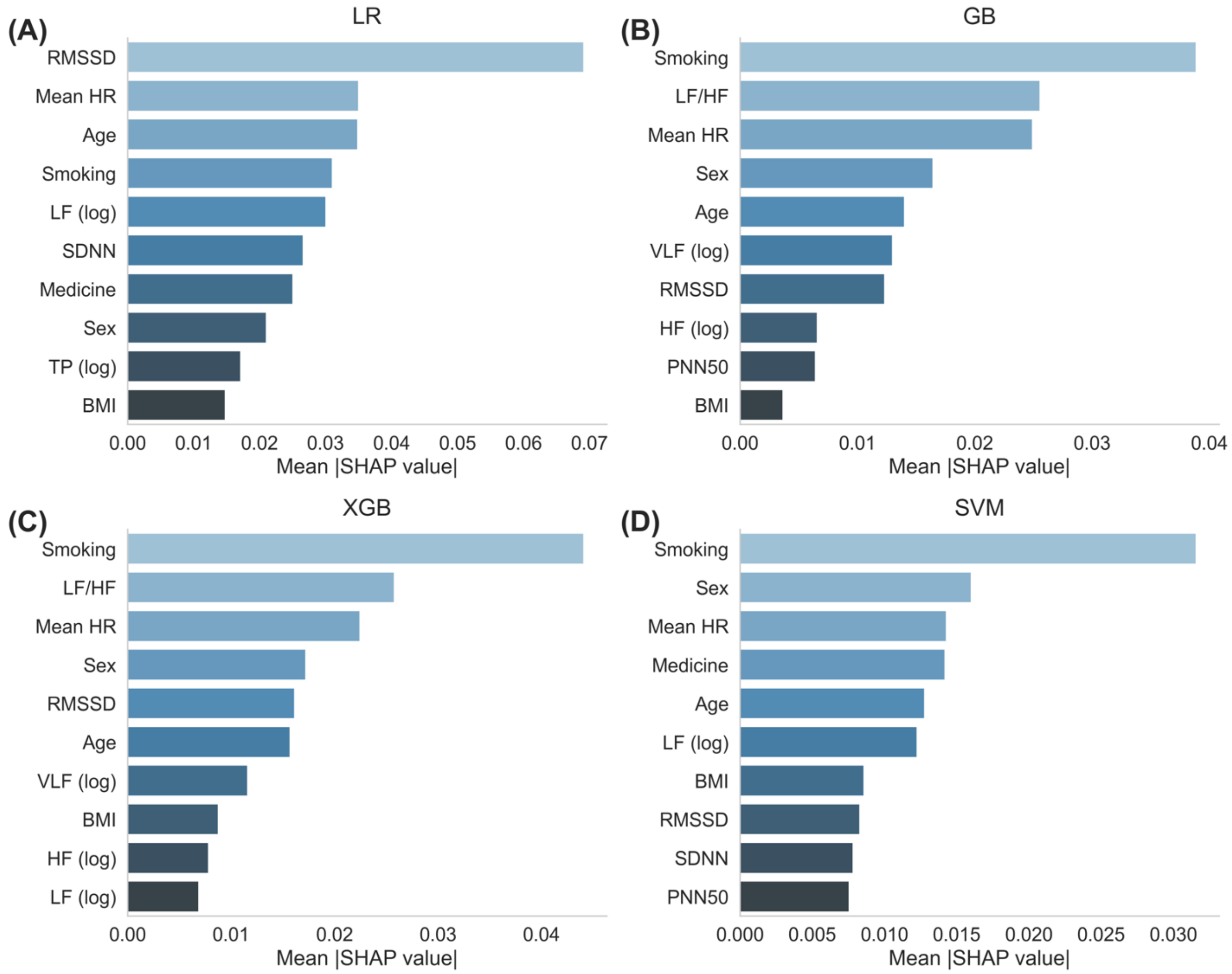

**Supplementary Table 1. Comparison of ECG-based and facial video-based HRV metrics in the MAHNOB-HCI dataset.**

| HRV Metric | ECG-based Mean | Video-based Mean | MAE (ms) | MAE (%) |
|---|---|---|---|---|
| HR (bpm) | 78.12 | 77.83 | 4.43 | 5.82% |
| SDNN (ms) | 114.53 | 114.4 | 7.58 | 9.23% |
| RMSSD (ms) | 52.91 | 53.63 | 7.4 | 22.76% |
| LF ($ms^2$) | 456.97 | 455.68 | 11.3 | 7.99% |
| HF ($ms^2$) | 509.55 | 509.76 | 8.21 | 8.84% |

MAE = Mean Absolute Error; HR = heart rate; SDNN = standard deviation of the normal-to-normal intervals; RMSSD = root mean square of successive RR interval differences; LF = low-frequency; HF = high-frequency
